# The role of inflammation in the prospective associations between early childhood sleep problems and ADHD at 10 years: Findings from a UK birth cohort study

**DOI:** 10.1101/2022.06.06.22276028

**Authors:** Isabel Morales Muñoz, Rachel Upthegrove, Kate Lawrence, Sandra Kooij, Alice M Gregory, Steven Marwaha

## Abstract

**Background:** Several underlying mechanisms potentially account for the link between sleep and attention deficit and hyperactivity disorder (ADHD), including inflammation. However, studies so far have been cross-sectional. We investigate (i) the association between early childhood sleep and probable ADHD diagnosis in childhood; and (ii) whether childhood circulating inflammatory markers mediate any associations.

**Methods and Findings:** Data from the Avon Longitudinal Study of Parents and Children (ALSPAC) were available for 7658 10-years-old children. Parent-reported sleep duration, night awakening frequency, and regular sleep routines were collected at 3.5 years. The Development and Wellbeing Assessment (DAWBA) was administered to capture children with clinically relevant ADHD symptoms, or probable ADHD diagnosis. Further, blood samples were collected at 9 years, from which two inflammatory markers were obtained [i.e. interleukin-6 (IL-6) and C-reactive protein (CRP)]. Logistic regressions were applied to investigate the associations between sleep variables at 3.5 years and probable ADHD diagnosis at 10 years. Further, path analysis was applied to examine the mediating role of inflammation at 9 years (i.e. as measured by CRP and IL-6) in the associations between early sleep and ADHD at 10 years. We found that less regular sleep routines (OR=0.51, 95%CI=0.28-0.93, p=0.029), shorter nighttime sleep (OR=0.70, 95%CI=0.56-0.89, p=0.004), and higher night awakening (OR=1.27, 95I%CI=1.06-1.52, p=0.009) at 3.5 years were associated with higher odds of probable ADHD at 10 years. Further, IL-6 at 9 years mediated the association between irregular sleep routines and ADHD (bias-corrected estimate, -0.002; p=0.005); and between night awakening and ADHD (bias-corrected estimate, 0.002; p=0.003).

**Conclusions:** Several sleep problems in early childhood constitute a risk factor for probable ADHD diagnosis at 10 years. These associations may be mediated by inflammation, as measured by IL-6. These results open a new research vista to the pathophysiology of ADHD and highlight sleep and inflammation as potential preventative targets for ADHD.

## Introduction

A good night of sleep is considered to be essential for every aspect of a child’s functioning (1). Further, identification of sleep problems in children is important because they are associated with physical, cognitive, and socio-emotional problems (2). For instance, sleep problems are commonly comorbid with neurodevelopmental conditions, including Attention Deficit and Hyperactivity Disorder (ADHD) (3). Systematic reviews indicate that the prevalence of ADHD in children and young people is between 2% and 7%, with an average of around 5% (4), and is characterized by attention difficulties, impulsivity and/or hyperactivity that interfere with social functioning and/or development (5). The aetiology of ADHD is multifactorial and includes both genetic and environmental factors (6,7)., One manifestation of this aetiology is disturbed sleep and there has been a dramatic increase in new research to better understand the association between disturbed sleep and ADHD (8).

Sleep disturbances are commonly comorbid with ADHD, and between 70% and 85% of children with ADHD experience problems with sleep (9). Sleep problems, such as fragmented sleep, bedtime resistance, and/or sleep-onset insomnia in early childhood have been found to precede ADHD symptoms. For instance, sleep disturbances at age 4 are associated with attention problems at age 15 (10). Other studies have revealed similar associations at earlier stages, with sleep problems between the ages of 2–4 years being associated with attention deficits at age 5 (11) or short sleep duration from 3-24 months associating with inattention at 5 years (12). These studies suggest that sleep problems in early childhood may be an initial symptom, risk indicator of later ADHD or a causal factor in the development of ADHD symptoms. However, most of these longitudinal studies have focused on ADHD symptomatology, rather than ADHD diagnosis. Further, it is still unclear whether all or only some specific sleep problems in early childhood precede the development of ADHD, although previous reviews and meta-analyses suggest that different sleep variables are associated with ADHD symptoms (13,14). Further, sleep problems in individuals with ADHD can result in significant functional impairments that affect mood, attention, behavior, and ultimately quality of life,(15) which supports the necessity of further investigating sleep problems in ADHD.

If sleep and ADHD are robustly and prospectively associated, the next step would be to understand the potential mechanisms by which sleep difficulties may increase the risk for ADHD. This could help us better understand the aetiology of poorer outcomes in people with ADHD and consequently design more targeted interventions. Among these potential candidates, inflammation has recently received some attention, and preliminary evidence suggests that inflammatory processes could contribute to the association between sleep and ADHD (16), although this requires further research consideration.

Sleep disturbances have been associated with altered levels of inflammatory cytokines in adults (17), but data in youth is still lacking. Two of the inflammatory markers that have received greatest attention in relation to sleep are interleukin-6 (IL-6) and C-reactive protein (CRP) (17). IL-6 is a pro-inflammatory cytokine produced in response to environmental stressors, infections and injuries. Dysregulated and persistent IL-6 production in certain cell populations leads to the development of various diseases (18). CRP is a pentameric protein synthesized by the liver, and its level rises in response to inflammation (19). A recent study reported that morning serum IL-6 associates with actigraphy-based night sleep efficiency and minutes spent awake in children and adolescents (20). Further, a recent study reported higher CRP and IL6 levels in children with ADHD, compared to healthy children (21,22). Overall, there is recent evidence to support the potential contributing role of inflammation in the association between sleep and ADHD in children. Currently, as far as we are aware there is no research which has specifically investigated the prospective association between inflammation, sleep and ADHD. Increased understanding here would allow us to further interrogate potential mechanisms underlying the links between disturbed sleep and ADHD and potentially better targeted early interventions in ADHD.

Therefore, the aims of this study were to investigate (i) the association between sleep variables (i.e., regular sleep routines, daytime sleep duration, nighttime sleep duration and night awakenings) in early childhood and probable ADHD diagnosis in childhood; and (ii) whether circulating markers of inflammation (i.e., CRP and IL-6) in childhood mediate the associations between sleep variables and later probable ADHD diagnosis. We hypothesized that shorter nighttime sleep duration would be the key sleep variable to be associated with later ADHD,(12) and that CRP and IL-6 would mediate the prospective association between sleep and ADHD.

## Methods and Materials

### Participants

The Avon Longitudinal Study of Parents and Children (ALSPAC) is a UK birth cohort study examining the determinants of development, health and disease during childhood and beyond (23,24). Pregnant women resident in Avon, UK with expected dates of delivery 1st April 1991 to 31st December 1992 were invited to take part in the study. The ALSPAC study website contains details of all the data available through a fully searchable data dictionary and variable search tool (http://www.bristol.ac.uk/alspac/researchers/our-data/). Further details of the ALSPAC cohort are provided in Supplementary material. Informed consent for the use of data collected via questionnaires and clinics was obtained from participants following the recommendations of the ALSPAC Ethics and Law Committee at the time. Ethical approval was obtained from the ALSPAC Law and Ethics committee and the local research ethics committees.

### Measures

#### Sleep assessment at 3.5 years old

Parent-reported sleep routines regularity, daytime sleep duration, nighttime sleep duration, and night awakenings frequency were assessed when the child was 3.5 years old. This time-point was focused upon based on previous findings that this time-point showed the strongest prospective associations with mental health problems in adolescence (25). Further, by the age of 3, children’s sleep-wake patterns are consolidated (26), which allows us to better differentiate sleep difficulties from normative developmental patterns. These sleep variables were collected using questionnaires, and the mothers were asked about their child’s bed and wake up times, night awakenings frequency, daytime sleep duration, and whether the child had regular sleep routines. Nighttime sleep duration was operationalized as the period between reported bedtime and morning wake time.

#### Probable ADHD diagnosis at 10 years old

Probable ADHD diagnosis at 10 years old was assessed via maternal ratings, using the well-validated Development and Well-Being Assessment interview (DAWBA) (27). Further details of the DAWBA are provided in Supplementary material. Considering that most cases with ADHD are diagnosed when children are 6 to 11 years old (28), we focused on ADHD at 10 years old to make sure that the majority of the potential cases with ADHD would be characterized. The DAWBA was administered via computer, generating the following ‘probability bands’ (i.e. levels of prediction of the probability of disorder for a DSM-IV diagnosis of ADHD, ranging from 0–very unlikely to 5–probable): 0:<0.1% probability of children in this band having the disorder; 1:∼0.5%; 2:∼3%; 3:∼15%; 4:∼50%; 5:>70%, respectively. The top two levels of the DAWBA bands were used as computer-generated DAWBA diagnoses, which was the measure selected for this study. This follows previous studies using the DAWBA bands, which were validated in 7,912 British children (7-19 years) and 1,364 Norwegian children (11-13 years), using clinician-rated DAWBA diagnoses as a gold standard (29). Briefly, the prevalence estimates of the computer-generated DAWBA diagnoses were of roughly comparable magnitude to the prevalence estimates from the clinician-generated diagnoses, but the estimates were not always very close. In contrast, the estimated effect sizes, significance levels and substantive conclusions regarding risk factor associations were very similar or identical. Therefore, the computer-generated DAWBA diagnoses may provide a useful alternative to clinician-rated diagnoses, when studying associations with risk factors, generating rough prevalence estimates or implementing routine mental health screening. Therefore, here we use the term “probable ADHD diagnosis”, instead of ADHD diagnosis.

#### Inflammatory markers at 9 years old

Blood samples were collected from non-fasting participants during clinic assessment at 9 years at around the same time of the day for each participant. Samples were immediately spun, frozen and stored at −80 °C. There was no evidence of freeze-thaw cycles during storage. High-sensitivity CRP (hsCRP) was measured at one time point at the same laboratory by automated particle-enhanced immunoturbidimetric assay (Roche UK, Welwyn Garden City, UK). Additionally, IL-6 was measured by single enzyme-linked immunosorbent assay (R&D Systems). All assay coefficients of variation were <5%. Data on IL-6 and CRP were available for 5071 and 5081 participants, respectively. In the total sample, CRP values ranged from 0.01 to 67.74 mg/L. Fifty-nine subjects had CRP levels >10 mg/L and were excluded from analysis due to the risk of confounding by acute inflammatory state (e.g. infection). IL-6 values ranged from 0.01 to 20.05 pg/ml and all values were kept for the analyses. Higher levels of IL-6 (18) and CRP (30) are both associated with higher probability of infection. IL-6 and CRP levels were log-transformed and standardized (Z-transformed), following previous research in the topic (31).

#### Confounders

Multiple family risk factors were assessed using the Family Adversity Index (FAI) during pregnancy, at 2 years, and at 4 years. The FAI comprises 18 items on early parenthood, housing conditions, maternal education, financial difficulties, parents’ relationship, family size, family major problems, maternal psychopathology, parents’ substance abuse, crime records, partner support and social network. Points were summed at each time point for a total FAI score across the 3 time points. The FAI total score has a range of 0–18, with higher scores indicating higher number of family adversities across early childhood. We included this variable as confounder, as adverse family environment is associated with ADHD (32).

Other socio-economic factors selected as covariates were child’s sex, preterm delivery, ethnicity (white vs non-white), maternal age when baby was born, and maternal socioeconomic status, which was measured using the Cambridge Social Interaction and Stratification Scale, which provides a total score (33). All these variables were selected as relevant confounders as there is a higher prevalence of ADHD in males (34), preterm children (35), white children (36), children with young mothers (37), and low socio-economic status (38).

### Statistical Analysis

As 50.3% of the original sample was lost to attrition by the 10-year follow-up, we conducted logistic regressions to identify significant predictors of attrition. Children lost due to attrition were more frequently born preterm, had lower birth weight, had shorter gestational age, had younger mothers, had higher scores in family adversity, and were more often of non-white ethnicity (see supplementary table S1). Using the variables associated with selective dropout as the predictors, we fitted a logistic regression model (nonresponse vs response outcome) to determine weights for each individual using the inverse probability of response. The regression coefficients from this model were used to determine probability weights for the covariates in the main analyses.

A multi-staged analysis plan was developed. We first ran weighted logistic regression analyses in SPSS-v27, to ascertain the unadjusted and adjusted associations between sleep variables at 3.5 years and computer-generated DAWBA diagnoses of ADHD at 10 years. In Model A, we tested unadjusted associations. In Model B, we controlled for sex, maternal age at birth, FAI, ethnicity, preterm delivery and maternal socio-economic group. Further, the four sleep variables were evaluated together in the same model, to account for the potential overlap between sleep patterns in childhood. Because this study involves exploratory analysis of multiple separate hypotheses, as opposed to repeated analyses of a single hypothesis, we did not adjust for multiple testing (39).

To examine the potential mediating role of inflammation at 9 years, mediation models were tested using path analysis in SPSS-Amos v27, with maximum likelihood estimation to test the effect of childhood sleep variables at 3.5 years on computer-generated DAWBA diagnoses of ADHD at 10 years, with inflammatory markers at 9 years as mediating variables. We also controlled for the potential interaction of the explanatory variables (i.e., sleep variables). IL-6 and CRP were tested separately in independent mediation models. We included as explanatory variables only those sleep variables with significant associations in Model B, from the regression analyses. We also controlled for sex, FAI and preterm delivery, as these were the significant covariates from the regression analyses. We used bootstrapped bias-corrected confidence intervals and p values for assessing the significance of the standardized direct, indirect and total effects. Missing data was dealt with using the full information maximum likelihood method.

## Results

Data were available on 7,769 participants whose mothers reported on ADHD information at 10 years. Table 1 shows the frequencies and descriptive values of sociodemographic, sleep and clinical variables. Further, a description of these variables in children with probable ADHD diagnosis, compared to non-ADHD are presented in Table 2.

**Table 1.** Descriptive variables of the sample at 10 years old

| **Socio-demographic factors** | **Frequencies** |  | **Descriptives** |  |
| --- | --- | --- | --- | --- |
|  | N | % | Mean | SD |
| Sex (boys / girls) | 3919 / 3850 | 50.4 / 49.6 | --- | --- |
| Ethnicity (white / non-whites) | 7077 / 121 | 98.3 / 1.7 | --- | --- |
| Birth weight, grammes | --- | --- | 3420.01 | 542.77 |
| FAI, total score | --- | --- | 3.93 | 4.01 |
| Maternal age when birth | --- | --- | 29.05 | 4.58 |
| Gestational age, weeks | --- | --- | 39.45 | 1.85 |
| Preterm delivery (Y/N) | 347 / 4169 | 7.7 / 92.3 | --- | --- |
| Maternal socio-economic status | --- | --- | 54.95 | 13.54 |
| **Sleep variables at 3.5 years** | N | % | Mean | SD |
| Regular sleep routines (Y/N) | 6395 / 465 | 93.2 / 6.8 | --- | --- |
| Daytime sleep duration, hours | --- | --- | 0.35 | 0.70 |
| Nighttime sleep duration, hours | --- | --- | 10.73 | 0.84 |
| Night awakenings per night, n | --- | --- | 0.57 | 0.82 |
| **ADHD** | N | % | Mean | SD |
| DAWBA probable ADHD diagnosis (Y/N) | 128 / 7641 | 1.6 / 98.4 | --- | --- |
| **Inflammatory markers at 9 years** | N | % | Mean | SD |
| CRP mg/L | --- | --- | 0.55 | 0.21 |
| IL-6 pg/ml | --- | --- | 1.28 | 1.57 |

ADHD = Attention deficit and hyperactivity disorder; FAI=Family adversity index; DAWBA = Development and Well-Being Assessment interview CRP=C-reactive protein; IL-6=Interleukin 6; CRP values ranged from 0.01 to 10 mg/L; and IL-6 values ranged from 0.01 to 20.05 pg/ml

**Table 2.** Differences in socio-demographic and sleep variables between children with and without probable ADHD diagnosis at 10 years

| Variables | Probable ADHD diagnosis at 10 years |  | p |
| --- | --- | --- | --- |
|  | Yes (n = 128) | No (n = 7641) |  |
|  | N (%) | N (%) |  |
| Sex |  |  | <0.001 |
| Male | 104 (2.7) | 3815 (97.3) |  |
| Female | 24 (0.6) | 3826 (99.4) |  |
| Preterm delivery |  |  | <0.001 |
| Yes | 15 (4.3) | 332 (95.7) |  |
| No | 70 (1.7) | 4099 (98.3) |  |
| Regular sleep routines 3.5 years |  |  | 0.004 |
| Yes | 96 (1.5) | 6299 (98.5) |  |
| No | 15 (3.2) | 450 (96.8) |  |
| Ethnicity |  |  | 0.951 |
| White | 112 (1.6) | 6965 (98.4) |  |
| Other | 2 (1.7) | 119 (98.3) |  |
|  | Mean (SD) | Mean (SD) | p |
| Maternal age when born (years) | 28.41 (5.41) | 29.06 (4.57) | 0.117 |
| Family Adversity score | 7.19 (5.47)* | 3.87 (3.96) | <0.001 |
| Gestational age (weeks) | 39.02 (1.92) | 39.45 (1.85) | 0.011 |
| Maternal socio-economic status | 49.91 (13.27) | 55.03 (13.53) | <0.001 |
| Birth weight, kg | 3.31 (0.55) | 3.42 (0.54) | 0.035 |
| Daytime sleep duration, hours | 0.52 (0.87) | 0.35 (0.70) | 0.010 |
| Nighttime sleep duration, hours | 10.83 (1.18) | 10.73 (0.83) | 0.187 |
| Night awakenings per night, n | 0.84 (1.22) | 0.57 (0.81) | 0.001 |
| CRP mg/L | 0.52 (0.86)* | 0.56 (1.02) | 0.786 |
| IL-6 | 1.36 (1.37)* | 1.28 (1.57) | 0.707 |

CRP=C-reactive protein; IL-6=Interleukin 6
\*Children with probable ADHD diagnosis at 10 years old reported significant higher score in Family Adversity Index (i.e. higher frequency of family adversities in early childhood) compared to children with no diagnosis of ADHD. There were not statistically significant differences between ADHD and non-ADHD children for CRP and IL-6 levels; however, children with probable ADHD diagnosis at 10 years presented slightly lower CRP levels at 9 years, while higher IL-6 levels at 9 years.

The associations from the logistic regressions between sleep variables at 3.5 years and computer-generated DAWBA diagnoses of ADHD at 10 years appear on Table 3. In Model A, the four sleep variables at 3.5 years old were significantly associated with probable ADHD diagnosis at 10 years. Furthermore, in Model B, when we controlled for the covariates, children with less regular sleep routines had higher odds of probable ADHD diagnosis at 10 years (OR=0.51, 95% CI=0.28-0.93, p=0.029), children with shorter nighttime sleep duration had higher odds of probable ADHD diagnosis (OR=0.70, 95% CI=0.56-0.89, p=0.004), and children with higher night awakening frequency had higher odds of probable ADHD diagnosis (OR=1.27,95I% CI=1.06-1.52, p=0.009).

**Table 3.** Logistic regression analyses between sleep problems at 3.5 years and probable ADHD diagnosis at 10 years

| Probable ADHD diagnosis at 10 years |  |  |  |  |  |  |
| --- | --- | --- | --- | --- | --- | --- |
|  | Unadjusted model (Model A) |  |  | Adjusted model (Model B) |  |  |
|  | OR | CI 95% | p | OR | CI 95% | p |
| Regular sleep routines, 3.5 years | **.35** | **.20 to .63** | **<.001** | **.51** | **.28 to .93** | **.029** |
| Daytime sleep duration, hours, 3.5 years | **1.38** | **1.08 to 1.76** | **.009** | 1.24 | .97 to 1.58 | .088 |
| Nighttime sleep duration, hours, 3.5 years | **.73** | **.57 to .92** | **.007** | **.70** | **.56 to .89** | **.004** |
| Night awakenings per night, 3.5 years | **1.29** | **1.09 to 1.53** | **.003** | **1.27** | **1.06 to 1.52** | **.009** |
| Sex | --- | --- | --- | **.29** | **.18 to .47** | **<.001** |
| Maternal age birth | --- | --- | --- | 1.01 | .97 to 1.05 | .712 |
| FAI | --- | --- | --- | **1.12** | **1.08 to 1.16** | **<.001** |
| Ethnicity | --- | --- | --- | 1.32 | .43 to 4.06 | .630 |
| Preterm | --- | --- | --- | **.30** | **.18 to .50** | **<.001** |
| Socio-economic group | --- | --- | --- | 1.02 | .94 to 1.12 | .584 |

OR=Odd ratio; CI=Confidence interval.
Ethnicity=white vs non-white.
FAI=Family adversity index; it comprises 18 items (i.e., long index) on early parenthood, housing conditions, maternal education, financial difficulties, parents' relationship, family size, family major problems, maternal psychopathology, parents' substance abuse, crime records, partner support and social network. The FAI was assessed during pregnancy (long index), at 2 years (long index), and at 4 years (short index). The short index excludes social, practical, and financial support. Points were summed at each time point for a total FAI score across the 3 time points.
Maternal socioeconomic status was measured using the Cambridge Social Interaction and Stratification Scale, which provides a total score

In relation to IL-6 at 9 years as potential mediator of the association between early childhood sleep variables and computer-generated DAWBA diagnoses of ADHD at 10 years, model fit indexes indicated excellent model fit (χ2=0.27, P=.60; RMSEA=0; CIF=1.00). Consistent with the adjusted logistic regression, higher night awakening frequency at 3.5 years was directly and significantly associated with probable ADHD diagnosis at 10 years (β=0.035, p<0.001). Direct associations are shown in Figure 1. Further, we observed an indirect effect of IL-6 at 9 years in the association between irregular sleep routines at 3.5 years and probable ADHD diagnosis at 10 years (bias-corrected estimate, -0.002; 95% CI=-0.003 to -0.001, p=0.005); and between higher night awakening frequency at 3.5 years and probable ADHD diagnosis at 10 years (bias-corrected estimate, 0.002; 95% CI=0.001 to 0.003, p=0.003) (see Table S2 in Supplement, for the indirect effects).

**Figure 1.**
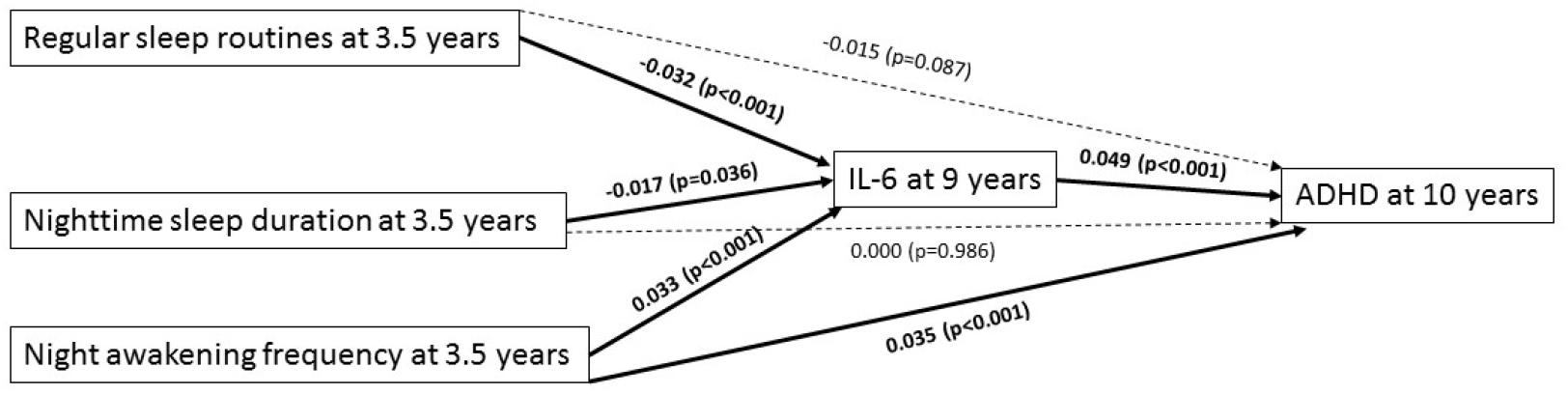
Path diagram showing the main direct associations, with IL-6 at 9 years as mediating factor. This figure shows only the direct associations of the independent, mediating and dependent variable. Sleep variables (i.e., regular sleep routines, nighttime sleep duration and night awakening frequency) at 3.5 years represent the independent variables; IL-6 at 9 years the mediating factor; and probable ADHD diagnosis at 10 years represents the outcome. The covariates also included in this path analyses were sex, family adversity and preterm delivery, due to the significant associations with the outcome found in the logistic regression model. Significant pathways are signified by solid arrows and nonsignificant pathways by dotted-dashed lines.

Concerning the potential mediating role of CRP at 9 years, we also demonstrated a good model fit (χ2=2.22, P=.14; RMSEA=0.009; CIF=1.00). Higher night awakening frequency at 3.5 years was the only early sleep problem which was directly and significantly associated with probable ADHD diagnosis at 10 years (β=0.036, p<0.001). Direct associations are shown in Figure 2. However, CRP at 9 years did not mediate any of the associations between early childhood sleep problems and probable ADHD diagnosis at 10 years (see Table S2 in Supplement, for the indirect effects).

**Figure 2.**
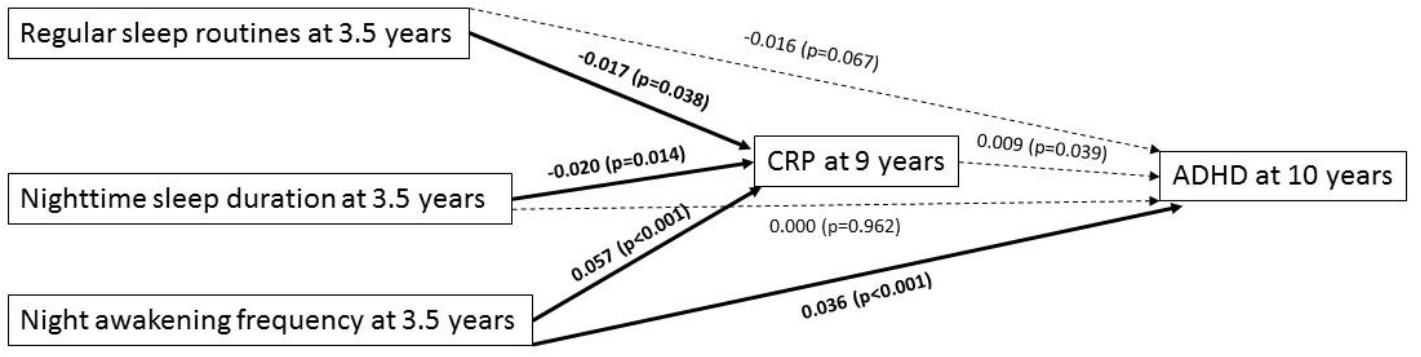
Path diagram showing the main direct associations, with CRP at 9 years as mediating factor. This figure shows only the direct associations of the independent, mediating and dependent variable. Sleep variables (i.e., regular sleep routines, nighttime sleep duration and night awakening frequency) at 3.5 years represent the independent variables; CRP at 9 years the mediating factor; and probable ADHD diagnosis at 10 years represents the outcome. The covariates also included in this path analyses were sex, family adversity and preterm delivery, due to the significant associations with the outcome found in the logistic regression model. Significant pathways are signified by solid arrows and nonsignificant pathways by dotted-dashed lines.

## Discussion

In a prospective well-established UK cohort study, we investigated whether several sleep variables at 3.5 years old were associated with the probable diagnosis of ADHD at 10 years old. To our knowledge, this is the first longitudinal study to examine whether inflammation mediates the association between early childhood sleep problems and subsequent probable ADHD diagnosis. First, we found that irregular sleep routines, shorter nighttime sleep duration and higher night awakening frequency at 3.5 years were all associated with probable ADHD diagnosis at 10 years. Second, we found that IL-6 at 9 years mediated the associations of both irregular sleep routines and higher frequency of night awakening at 3.5 years with ADHD at 10 years. In contrast, CRP at 9 years did not mediate any of the associations between early childhood sleep problems and probable ADHD diagnosis at 10 years.

We found that several parent-reported sleep problems in early childhood were prospectively associated with probable ADHD diagnosis at 10 years. This is consistent with previous research supporting the notion that sleep problems in early childhood might constitute an important risk factor for developing ADHD later in childhood. For instance, parent-reported short sleep duration at 5-6 years old is prospectively associated with ADHD symptoms at 8-9 years old (40). Furthermore, these prospective associations with short sleep duration have been also reported at earlier ages, with shorter sleep duration from 3-to-24 months being associated with inattention at 5 years (12). These prospective associations continue into adolescence, with recent evidence suggesting that several sleep problems in early childhood, including insomnia, restless leg syndrome and frequent snoring (41), as well as difficulty going to sleep, nightmares and restless sleep predict ADHD symptoms in adolescence (42). Our findings contribute to the existing evidence by highlighting that specific sleep problems, including short sleep length and sleep fragmentation in early childhood are also related to probable ADHD diagnosis at 10 years old. Further, irregular sleep routines at 3.5 years were also prospectively related to probable ADHD diagnosis at 10 years, suggesting that a lack of routine is also an important factor to consider with regards to ADHD, as has been previously reported (43).

Several underlying mechanisms have been suggested to account for the link between sleep and ADHD. For example, sleep deprivation can impact the prefrontal cortex (44), and weaker function and structure of prefrontal cortex circuits associate with ADHD (45). Further, dopaminergic activity is associated with ADHD symptomology (46), and is also implicated in the regulation of sleep and waking (47). In this study, we focused on the two most commonly assayed inflammatory markers (i.e. IL-6 and CRP) as potential mediators of the prospective associations between sleep and ADHD in childhood, due to a possible role of inflammation in ADHD pathogenesis (48).

Our initial hypothesis that both markers of inflammation would mediate this association was partially supported. We found that IL-6, but not CRP, at 9 years, mediated the associations between two of the sleep variables tested at 3.5 years (i.e., irregular sleep routines and frequent night awakening) with ADHD at 10 years. The impacts of IL-6 and CRP on subsequent outcomes, including mental health and brain development may differ. For instance, in a longitudinal study, higher levels of IL-6 in childhood, but not CRP, were associated with an increased risk of developing depression and psychosis in young adulthood (49). Similarly, IL-6 at 9 years, but not CRP, was prospectively associated with diurnal mood variation, concentration difficulties, fatigue and sleep disturbances at 18 years old (50), and with increased risk of hypomanic symptoms by age 22 years (51). A recent study also reported that genetically predicted IL-6, but not CRP, was significantly associated with gray matter volume (especially in the middle temporal gyrus and fusiform gyrus), and with cortical thickness (mainly in the superior frontal gyrus) (52), which are areas implicated in ADHD.

So far, it is still unknown why IL-6 but not CRP might play a greater role in the development of subsequent mental health problems. One potential explanation might be found in the different physiological actions associated with each of these inflammatory markers (53). However, it is still unknown how this might lead to different adverse outcomes and under which circumstances. Therefore, further research to understand and compare the potential distinct role of IL-6 and CRP in the development of mental disorders is needed.

This study has several strengths, including the large population-based sample size, the longitudinal design, and the inclusion of several sleep variables in early childhood. There are also some limitations. First, other potential contributing factors, such as depression, cognition, social interactions, obstetric complications, viral infections, smoking or body mass index were left unexplored. Second, DAWBA probable ADHD at 10 years was only reported by parents, and thus parental perceptions and/or assumptions could be constituting a bias of these assessments. Future studies should consider self-reported and/or teacher-reported assessments, in addition to clinical interviews. Similarly, sleep was reported by the parents, and thus future studies should focus on objective measures of sleep. Third, the time-points included in this study were partially determined by the availability of the data collected in the ALSPAC. Future studies should also consider other developmental stages to investigate this topic. Fourth, as might be expected in a long-term population cohort study, the attrition rate was significant. However, we used procedures to ensure representativeness of our results. Finally, the prevalence of children with probable ADHD diagnosis using the top two levels of the DABWA-bands as computer-generated DAWBA diagnoses in this study was considerably smaller (i.e., 1.6%) than the prevalence reported by previous studies (e.g., around 5%). These discrepancies in the prevalence might be related to the different methodology used to assess ADHD and the specific time points investigated. However, the lower prevalence detected using the DAWBA in our study supports the assumption that the DAWBA represents clinically relevant ADHD cases, and that only includes those children with probable ADHD diagnosis.

## Conclusions

Our findings showed that several sleep variables in early childhood, including shorter nighttime sleep duration, higher night awakening and more irregular sleep routines at 3.5 years were associated with probable ADHD diagnosis at 10 years. Further, IL-6 at 9 years, but not CRP at 9 years mediated the associations between early sleep problems (i.e., higher night awakening and more irregular sleep routines) and probable ADHD diagnosis at 10 years. These findings support the relevant role of sleep problems at early stages of life as a risk factor for developing ADHD. In addition, a non-resolving pro-inflammatory mechanism might be a contributory pathway explaining why sleep problems in early childhood are linked to subsequent ADHD, and thus provides further support to the role of inflammation in mechanistic pathways to neurodevelopmental disorders, including ADHD. These results highlight the potential of future preventative interventions in ADHD, with the novel target of sleep and inflammation.

## Data Availability

The ALSPAC data and other materials/codes used for this study can be provided by ALSPAC pending scientific review and a completed material transfer agreement. Requests for the ALSPAC data should be submitted to.

## Contributors

IMM, SM, RU, AG and KL, designed the study. IMM was primarily responsible for data analysis and writing of the article. SM, RU, SK, AG and KL, reviewed and contributed critically to the writing of the article. All authors reviewed and contributed to the manuscript and approved the final version of the manuscript.

## Acknowledgments

The UK Medical Research Council and Wellcome (Grant ref: 217065/Z/19/Z) and the University of Bristol provide core support for ALSPAC. We are extremely grateful to all the families who took part in this study, the midwives for their help in recruiting them, and the whole ALSPAC team, which includes interviewers, computer and laboratory technicians, clerical workers, research scientists, volunteers and managers.

## Supplementary material

### Further details of the ALSPAC cohort

The initial number of pregnancies enrolled was 14,541 (for these at least one questionnaire was returned, or a “Children in Focus” clinic had been attended by 19/07/99). Of these initial pregnancies, there was a total of 14676 foetuses, resulting in 14062 live births and 13988 children who were alive at 1 year of age. When the oldest children were approximately 7 years of age, an attempt was made to bolster the initial sample with eligible cases who had failed to join the study originally. As a result, in our study, as some variables were collected from the age of seven onwards there were data available for more than the 14541 pregnancies mentioned above. Informed consent for the use of data collected via questionnaires and clinics was obtained from participants following the recommendations of the ALSPAC Ethics and Law Committee at the time. Ethical approval was obtained from the ALSPAC Law and Ethics committee and the local research ethics committees.

### Further details of the Development and Well-Being Assessment (DAWBA)

The DAWBA is a package of interviews, questionnaires and rating techniques designed to generate ICD-10 and DSM-IV or DSM-5 psychiatric diagnoses about 2-17 years old. The DAWBA includes a mix of ‘closed’/structured questions and open-ended questions, where respondents describe their difficulties in their own words. The full DAWBA package covers the following diagnoses: Separation anxiety, Specific phobia, Social phobia, Panic disorder/agoraphobia, Post-traumatic stress disorder, Obsessive compulsive disorder, Generalized anxiety disorder, Body dysmorphic disorder, Disruptive mood dysregulation disorder, Major depression, ADHD/hyperkinesis, Oppositional defiant disorder, Conduct disorder, Eating disorders, including anorexia, bulimia and binge eating, Autism spectrum disorders, Tic disorders, including Tourette syndrome, and Bipolar disorders. For each of these disorders, the interview asks about all the symptoms, and other criteria needed for an operationalized diagnosis according to both DSM-IV (American Psychiatric Association, 1994) and the research diagnostic version of ICD-10 (World Health Organisation, 1994). Panic disorder, agoraphobia, autistic disorders, eating disorders, tic disorders, and any other concerns are covered more briefly, with clinical diagnoses of these disorders being correspondingly more dependent on rating the open-ended transcript. The time frame of the interview is the present and the recent past. For many disorders, the ICD-10 and DSM-IV diagnostic criteria stipulate that the symptoms need to have persisted for a specified number of months, e.g. a minimum of 6 months for hyperactivity, oppositional-defiant disorder, and generalized anxiety disorders. In these instances, the relevant section of the DAWBA interview focuses on the child’s symptoms over this stipulated period. The time frame is longest for conduct disorder (since DSM-IV criteria include the number of relevant behaviours displayed over the previous 12 months), and shortest for most of the emotional disorders, where the focus is on the last month, in line with previous recommendations (Shaffer et al., 1996).

**Table S2.** Differences in socio-demographic variables between non-participating and participating subjects in the study at 10 years old

|  | Non-participating group in the study |  | Participating group in the study |  | Non-participating versus participating |  |
| --- | --- | --- | --- | --- | --- | --- |
|  | *Mean* | *SD* | *Mean* | *SD* | *OR (95% CI)* | *p* |
| Maternal age when born | 26.81 | 5.11 | 29.05 | 4.58 | 1.10 (1.09 to 1.11) | <0.001 |
| Gestational age | 37.24 | 7.46 | 39.45 | 1.85 | 1.14 (1.10 to 1.13) | <0.001 |
| Birth weight, kg | 3.34 | 0.62 | 3.42 | 0.54 | 1.28 (1.21 to 1.35) | <0.001 |
| Family Adversity score | 5.34 | 4.75 | 3.93 | 4.01 | 0.93 (0.92 to 0.94) | <0.001 |
|  | Non-participating group in the study |  | Participating group in the study |  |  |  |
|  | *N* | *%* | *N* | *%* |  |  |
| Sex |  |  |  |  | 0.94 (0.88 to 1.01) | 0.077 |
| Male / Female | 3772 / 3498 | 51.9 / 48.1 | 3919 / 3850 | 50.4 / 49.6 |  |  |
| Ethnicity |  |  |  |  | 0.42 (0.33 to 0.52) | <0.001 |
| White / Other | 4985 / 205 | 96.1 / 3.9 | 7077 / 1.7 | 98.3 / 1.7 |  |  |
| Preterm delivery |  |  |  |  | 0.56 (0.49 to 0.65) | <0.001 |
| Yes / No | 509 / 3443 | 12.9 / 87.1 | 347 / 4169 | 7.7 / 92.3 |  |  |

The individuals associated with attrition at 10 years were more frequently born preterm, had lower weight at birth, had shorter gestational age, had younger mothers at birth, had higher scores in family adversity, and were more often of non-white ethnicity.

**Table S2.** Bootstrapped bias-corrected confidence intervals and p values for the hypothesized indirect pathways to probable ADHD diagnosis at 10 years with IL6 at 9 years, and CRP at 9 years as mediators

|  | Probable ADHD diagnosis at 10 years<br>Via IL-6 at 9 years |  |  | Probable ADHD diagnosis at 10 years<br>Via CRP at 9 years |  |  |
| --- | --- | --- | --- | --- | --- | --- |
|  | Lower Bound | Upper Bound | p | Lower Bound | Upper Bound | p |
| Nigh awakening frequency 3.5 years | **0.001** | **0.003** | **0.003** | -0.001 | 0.000 | 0.329 |
| Regular sleep routines 3.5 years | **-0.003** | **-0.001** | **0.005** | -0.001 | 0.000 | 0.268 |
| Night sleep duration 3.5 years | -0.002 | 0.000 | 0.093 | 0.000 | 0.001 | 0.365 |

IL-6=Interleukin 6; CRP=C-reactive protein

